# Trends in mechanical circulatory support use and outcomes of patients with cardiogenic shock in Japan, 2010-2020: a nationwide inpatient database study

**DOI:** 10.1101/2023.02.07.23285628

**Authors:** Yuji Nishimoto, Hiroyuki Ohbe, Hiroki Matsui, Jun Nakata, Toru Takiguchi, Mikio Nakajima, Yusuke Sasabuchi, Yukihito Sato, Tetsuya Watanabe, Takahisa Yamada, Masatake Fukunami, Hideo Yasunaga

**Affiliations:** Division of Cardiology, Osaka General Medical Center, Osaka, Japan; Department of Clinical Epidemiology and Health Economics, School of Public Health, The University of Tokyo, Tokyo, Japan; Division of Cardiovascular Intensive Care, Nippon Medical School, Tokyo, Japan; Department of Emergency and Critical Care Medicine, Nippon Medical School, Tokyo, Japan; Emergency Life-Saving Technique Academy of Tokyo, Foundation for Ambulance Service Development, Tokyo, Japan; Data Science Center, Jichi Medical University, Tochigi, Japan; Department of Cardiology, Hyogo Prefectural Amagasaki General Medical Center, Amagasaki, Japan

**Keywords:** cardiogenic shock, mechanical circulatory support, extracorporeal membrane oxygenation, intra-aortic balloon pumping, Impella, epidemiology

## Abstract

**Background:** Little is known about the impact of the downgrade of guideline recommendations for intra-aortic balloon pump (IABP) use and the approval of the Impella in Japan, where IABPs have been enthusiastically used. This study aimed to describe the annual trends in the mechanical circulatory support (MCS) use and outcomes in patients with cardiogenic shock (CS) requiring MCS.

**Methods:** Using the Japanese Diagnosis Procedure Combination database from July 2010 to March 2021, we identified inpatients with CS requiring MCS. The patients were stratified into three groups: (i) IABP alone, (ii) Impella alone, and (iii) extracorporeal membrane oxygenation (ECMO) regardless of IABP or Impella use. The patient characteristics and outcomes were reported by the fiscal year.

**Results:** Of the 160,559 eligible patients, 117,599 (73.2%) used IABP alone, 1,465 (0.9%) Impella alone, and 41,495 (25.8%) ECMO. The prevalence of an IABP alone significantly decreased from 80.5% in 2010 to 65.3% in 2020 (P for trend <0.001), whereas the prevalence of an Impella alone significantly increased from 0.0% to 5.0% as well as ECMO from 19.5% to 29.6% (P for trend <0.001 for both). In-hospital mortality significantly increased from 29.3% in 2010 to 32.6% in 2020 in the overall patients with CS requiring MCS, but significantly decreased in those requiring ECMO from 73.7% to 64.1% (P for trend <0.001 for both).

**Conclusions:** There were significant annual changes in the patterns of MCS use and clinical outcomes in patients with CS requiring MCS.

## Introduction

Despite the widespread use of primary percutaneous coronary intervention (PCI) for ST-elevation myocardial infarction (STEMI) and advances in critical care, early mortality of cardiogenic shock (CS) remains high at 30–50% over the past two decades.^1,2^ In addition, the incidence of CS has increased due to an improved diagnostic accuracy, better access to health care, and increased patients with non-STEMI as well as non-ischemic etiologies.^3–6^

Given that previous studies showed no survival benefit of an intra-aortic balloon pump (IABP) for patients with acute myocardial infarction regardless of concomitant CS,^7–9^ recent guidelines no longer recommend routine IABP use in this context from 2013 in the United States, 2014 in Europe, and 2018 in Japan.^10–12^ Consequently, the use of an IABP has drastically decreased in the United States and Europe, whereas, an increasing trend has been shown in the use of an Impella, a new percutaneous ventricular assist device.^13,14^ Since October 2017, the Impella has been available in Japan, where IABPs have been used with great enthusiasm.^15,16^ However, it is unclear whether these major changes in CS practice have affected the patterns of mechanical circulatory support (MCS) use and clinical outcomes in Japan.

Capturing the trends in CS practice following changes in the guideline recommendations and approval of the new MCS device is important to understand the current status of CS practice and to establish evidence-based medicine in the future. Therefore, the present study aimed to describe the prevalence of MCS use and outcomes in patients with CS requiring MCS and their trends over time, using a nationwide inpatient database in Japan.

## Methods

### Design and Ethical statement

This was a retrospective cohort study using an inpatient administrative database, and the study conformed to the RECORD reporting guidelines.^17^ This study was conducted in accordance with the amended Declaration of Helsinki and was approved by the Institutional Review Board of The University of Tokyo (approval number, 3501-(5); 19 May 2021). Because the data were anonymous, the Institutional Review Board waived the requirement for informed consent.

### Data source

We used the Japanese Diagnosis Procedure Combination inpatient database, which contained administrative claims data and discharge abstracts from more than 1,500 acute care hospitals and covered approximately 90% of all tertiary emergency hospitals in Japan.^18^ The database includes the following patient-level data for all hospitalizations: age, sex, diagnoses (main diagnosis, admission-precipitating diagnosis, most resource-consuming diagnosis, second-most resource-consuming diagnosis, comorbidities present at admission, and complications arising after admission) recorded with the *International Classification of Diseases, 10th Revision (ICD-10)* codes, daily procedures recorded using Japanese medical procedure codes, daily drug administration, and discharge status.^18^ A previous validation study showed that the specificity of the recorded diagnoses in the database exceeded 96%, the sensitivity of the diagnoses ranged from 50% to 80%, and the specificity and sensitivity of the procedures both exceeded 90%.^19^

### Study population

Using the data from July 2010 to March 2021, we included inpatients who required MCS devices (IABP, Impella, or ECMO) during the hospitalization. Of those, we excluded the patients without a primary diagnosis of cardiovascular disease as defined by the ICD-10 codes listed in **Supplemental Table S1**. The primary diagnosis was defined when it appeared as a main diagnosis, admission-precipitating diagnosis, most resource-consuming diagnosis, or second most resource-consuming diagnosis.^19^

The eligible patients were stratified into three groups according to the MCS devices during the hospitalization: (i) IABP alone, (ii) Impella alone, and (iii) ECMO. The ECMO group included patients who received ECMO regardless of IABP or Impella use during the hospitalization.

### Variables and outcomes

The variables included the number of hospitals, hospital volume, age, sex, body mass index at admission, Japan Coma Scale at admission,^20^ ambulance use, primary diagnoses (acute coronary syndrome [ACS], aortic disease, valve disease, heart failure, cardiac arrest, ventricular tachycardia or fibrillation, pulmonary embolism, myocarditis, or cardiomyopathy), out-of-hospital cardiac arrest (OHCA) (admission-precipitating diagnosis), and treatments during the hospitalization (PCI, coronary artery bypass grafting [CABG], surgical valve procedures [repair or replacement], percutaneous valve procedures [repair, replacement or balloon dilatation], ventricular assist device, or heart transplantation).

The primary outcome was in-hospital mortality. The secondary outcomes were the length of MCS, length of hospital stay, total hospitalization cost, major bleeding, ischemic stroke, continuous renal replacement therapy, and total blood transfusion volume during the hospitalization. Major bleeding was defined as the presence of either intracranial bleeding (ICD-10 code: I61), intraspinal bleeding (G951), pericardial hematomas (I312), intra-abdominal or retroperitoneal hematomas (K661), intra-articular bleeding (M250), intraocular bleeding (H448), or compartment syndrome (M622), which was in accordance with the International Society of Thrombosis and Haemostasis definition.^21^

### Statistical analysis

Continuous variables were presented as the mean and standard deviation (SD), and categorical variables were presented as the number and percentage. To evaluate the trends by the fiscal year, we performed Cochran-Armitage tests for the binary variables and Jonckheere-Terpstra tests for the continuous variables. The trends in the patient characteristics and outcomes were assessed among overall patients with CS requiring MCS and those requiring each MCS device. The age and body mass index had missing values as shown in **Supplementary Table S2**. We considered all reported p-values as two-sided and a p<0.05 as statistically significant. All analyses were performed using STATA/SE 17.0 software (StataCorp).

## Results

### Patient characteristics and outcomes

During the study period, we identified 160,559 patients with CS requiring MCS during the hospitalization from 1,107 hospitals. Of those, 117,599 (73.2%) were identified as the IABP alone group, 1,465 (0.9%) as the Impella alone group, and the remaining 41,495 (25.8%) as the ECMO group. Of the overall 1,107 hospitals, IABP alone was used in 1,096 (99.0%), Impella alone in 120 (10.8%), and ECMO in 831 (75.1%) (**Table 1**). The patients in the IABP alone group were the oldest and those in the ECMO group were the youngest. The prevalence of patients with ACS was 78.6%, 63.0%, and 44.8% in the IABP alone, Impella alone, and ECMO groups, respectively, while those with OHCA was 3.4%, 7.3%, and 23.6% and those with myocarditis was 0.8%, 9.7%, and 4.0%. More than half of the patients in the IABP alone and Impella alone groups underwent PCI. Of the patients in the IABP alone group, 22.3% received CABG and 8.2% received surgical valve procedures. In-hospital mortality was 17.4%, 38.0%, and 68.3% in the IABP alone, Impella alone, and ECMO groups, respectively. The length of hospital stay was 32.6, 36.6, and 29.4 days and the total hospitalization cost was 4,522, 9,098, and 5,840 thousand yen in the IABP alone, Impella alone, and ECMO groups, respectively. All of major bleeding, ischemic stroke, and continuous renal replacement therapy were most prevalent in the Impella alone group and least prevalent in the IABP alone group.

**Table 1.** Patient characteristics.

|  | Overall<br>(*n* = 160,559) | IABP alone<br>(*n* = 117,599) | Impella alone<br>(*n* = 1,465) | ECMO<br>(*n* = 41,495) |
| --- | --- | --- | --- | --- |
| Number of hospitals, *n* | 1,107 | 1,096 | 120 | 831 |
| Hospital volume, median (IQR) | 66 (17-196) | 55 (15-147) | 9 (4-16) | 22 (7-64) |
| Age, years, mean (SD) | 69 (14) | 70 (12) | 66 (15) | 64 (16) |
| Men, *n* (%) | 117,013 (72.9) | 85,921 (73.1) | 1,108 (75.6) | 29,984 (72.3) |
| Body mass index on the day of admission, kg/m<sup>2</sup>, mean (SD) | 23.5 (4.7) | 23.4 (4.5) | 23.4 (4.1) | 23.9 (5.4) |
| Japan Coma Scale on the day of admission, *n* (%) |  |  |  |  |
| 0 (alert) | 112,448 (70.0) | 92,496 (78.7) | 856 (58.4) | 19,096 (46.0) |
| 1-3 (dizzy) | 13,680 (8.5) | 10,502 (8.9) | 163 (11.1) | 3,015 (7.3) |
| 10-30 (somnolent) | 5,054 (3.1) | 3,514 (3.0) | 74 (5.1) | 1,466 (3.5) |
| 100-300 (coma) | 29,377 (18.3) | 11,087 (9.4) | 372 (25.4) | 17,918 (43.2) |
| Ambulance use, *n* (%) | 100,028 (62.3) | 69,220 (58.9) | 1,113 (76.0) | 29,695 (71.6) |
| Primary diagnosis, *n* (%) |  |  |  |  |
| Acute coronary syndrome | 111,971 (69.7) | 92,476 (78.6) | 923 (63.0) | 18,572 (44.8) |
| Aortic disease | 4,762 (3.0) | 954 (0.8) | 17 (1.2) | 3,791 (9.1) |
| Valve disease | 12,819 (8.0) | 9,358 (8.0) | 86 (5.9) | 3,375 (8.1) |
| Heart failure | 24,186 (15.1) | 19,509 (16.6) | 315 (21.5) | 4,362 (10.5) |
| Ventricular tachycardia or fibrillation | 6,488 (4.0) | 2,365 (2.0) | 62 (4.2) | 4,061 (9.8) |
| Pulmonary embolism | 2,335 (1.5) | 126 (0.1) | 0 (0.0) | 2,209 (5.3) |
| Myocarditis | 2,741 (1.7) | 926 (0.8) | 142 (9.7) | 1,673 (4.0) |
| Cardiomyopathy | 8,541 (5.3) | 6,632 (5.6) | 134 (9.1) | 1,775 (4.3) |
| Cardiac arrest | 16,834 (10.5) | 4,770 (4.1) | 133 (9.1) | 11,931 (28.8) |
| Out-of-hospital cardiac arrest | 13,957 (8.7) | 4,038 (3.4) | 107 (7.3) | 9,812 (23.6) |
| Treatments, *n* (%) |  |  |  |  |
| Percutaneous coronary intervention | 89,507 (55.6) | 72,367 (61.5) | 899 (61.4) | 16,241 (39.1) |
| Coronary artery bypass grafting | 29,899 (18.6) | 26,170 (22.3) | 94 (6.4) | 3,635 (8.8) |
| Surgical valve procedures | 12,028 (7.5) | 9,625 (8.2) | 74 (5.1) | 2,329 (5.6) |
| Percutaneous valve procedures | 1,953 (1.2) | 843 (0.7) | 34 (2.3) | 1,076 (2.6) |
| Ventricular assist device | 976 (0.6) | 464 (0.4) | 23 (1.6) | 489 (1.2) |
| Heart transplantation | 57 (0.04) | 31 (0.03) | 0 (0.0) | 26 (0.1) |
| Outcomes |  |  |  |  |
| In-hospital mortality, *n* (%) | 49,407 (30.8) | 20,513 (17.4) | 556 (38.0) | 28,338 (68.3) |
| Length of MCS, days, mean (SD) | 4.0 (6.1) | 3.5 (4.0) | 7.2 (7.3) | 5.4 (9.8) |
| Length of hospital stay, days, mean (SD) | 31.8 (45.8) | 32.6 (40.5) | 36.6 (37.8) | 29.4 (58.6) |
| Total hospitalization cost, ×10<sup>3</sup> yen, mean (SD) | 4,905 (5,096) | 4,522 (3,917) | 9,098 (5,754) | 5,840 (7,344) |
| Major bleeding in a critical area or organ, *n* (%) | 1,110 (0.7) | 445 (0.4) | 32 (2.2) | 633 (1.5) |
| Ischemic stroke, *n* (%) | 3,908 (2.4) | 2,735 (2.3) | 60 (4.1) | 1,113 (2.7) |
| Continuous renal replacement therapy, *n* (%) | 31,265 (19.5) | 17,145 (14.6) | 509 (34.7) | 13,611 (32.8) |
| Blood transfusions, ml, mean (SD) |  |  |  |  |
| Red blood cells | 1,743 (23,815) | 1,037 (15,824) | 3,062 (3,947) | 3,699 (38,459) |
| Fresh-frozen plasma | 1,148 (27,702) | 632 (10,578) | 1,670 (3,266) | 2,592 (51,469) |
| Platelet concentrate | 271 (4,260) | 168 (3,334) | 511 (1,066) | 555 (6,209) |

ECMO, extracorporeal membrane oxygenation; IABP, intra-aortic balloon pumping; IQR, interquartile range; SD, standard deviation

### Trends in MCS use and outcomes

The prevalence of an IABP alone significantly decreased from 80.5% in 2010 to 65.3% in 2020 (P for trend <0.001) (**Figure 1**). In contrast, the prevalence of an Impella alone significantly increased from 0.0% in 2010 to 5.0% in 2020, and the prevalence of ECMO significantly increased from 19.5% in 2010 to 29.6% in 2020.

**Figure 1.**
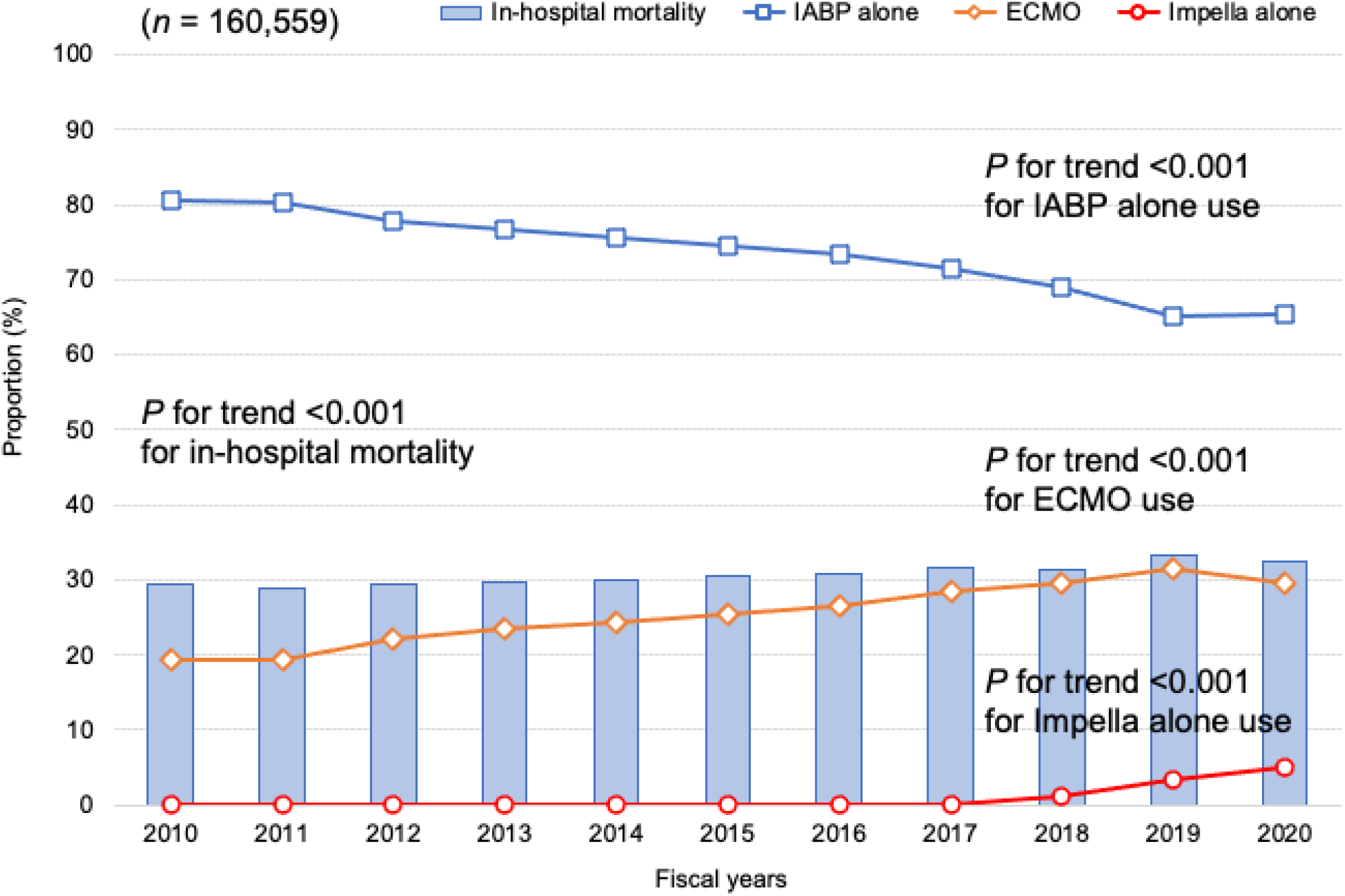
Annual proportion of in-hospital mortality and prevalence of IABP alone, Impella alone, and ECMO use in the overall patients with cardiogenic shock ECMO, extracorporeal membrane oxygenation; IABP, intra-aortic balloon pump

Among 160,559 patients with CS requiring MCS, the prevalence of ACS significantly decreased from 74.7% in 2010 to 65.1% in 2020, whereas the prevalence of valve disease and OHCA significantly increased from 5.6% and 7.2% in 2010 to 9.9% and 9.4% in 2020, respectively (**Table 2**). There was no significant change in the prevalence of heart failure. The prevalence of patients who underwent PCI and CABG significantly decreased, whereas the prevalence of patients who underwent percutaneous valve procedures significantly increased. In-hospital mortality significantly increased from 29.3% in 2010 to 32.6% in 2020 (**Figure 1** and **Table 2**).

**Table 2.** Patient characteristics and outcomes on an annual basis in overall patients requiring MCS.

|  | Fiscal year |  |  |  |  |  |  |  |  |  |  | *P* for trend |
| --- | --- | --- | --- | --- | --- | --- | --- | --- | --- | --- | --- | --- |
|  | 2010<br>(*n* = 8,712) | 2011<br>(*n* = 13,146) | 2012<br>(*n* = 14,213) | 2013<br>(*n* = 13,914) | 2014<br>(*n* = 15,849) | 2015<br>(*n* = 15,917) | 2016<br>(*n* = 16,585) | 2017<br>(*n* = 16,614) | 2018<br>(*n* = 15,939) | 2019<br>(*n* = 14,937) | 2020<br>(*n* = 14,733) |  |
| Age, years, mean (SD) | 69 (13) | 69 (13) | 68 (13) | 68 (13) | 68 (14) | 69 (13) | 69 (14) | 69 (14) | 69 (14) | 69 (14) | 70 (14) | 0.004 |
| Men, *n* (%) | 6,354 (72.9) | 9,574 (72.8) | 10,216 (71.9) | 10,206 (73.4) | 11,622 (73.3) | 11,539 (72.5) | 12,174 (73.4) | 12,076 (72.7) | 11,622 (72.9) | 10,890 (72.9) | 10,740 (72.9) | 0.58 |
| Body mass index on the day of admission, kg/m<sup>2</sup>, mean (SD) | 23.5 (7.7) | 23.4 (3.8) | 23.3 (3.8) | 23.4 (3.8) | 23.4 (4.0) | 23.5 (4.0) | 23.6 (7.2) | 23.6 (4.1) | 23.6 (4.2) | 23.6 (4.2) | 23.7 (4.6) | 0.001 |
| Primary diagnosis, *n* (%) |  |  |  |  |  |  |  |  |  |  |  |  |
| Acute coronary syndrome | 6,512 (74.7) | 9,835 (74.8) | 10,151 (71.4) | 10,041 (72.2) | 11,342 (71.6) | 11,319 (71.1) | 11,574 (69.8) | 11,251 (67.7) | 10,700 (67.1) | 9,656 (64.6) | 9,590 (65.1) | <0.001 |
| Valve disease | 486 (5.6) | 813 (6.2) | 1,044 (7.3) | 971 (7.0) | 1,196 (7.5) | 1,209 (7.6) | 1,399 (8.4) | 1,488 (9.0) | 1,383 (8.7) | 1,373 (9.2) | 1,457 (9.9) | <0.001 |
| Heart failure | 1,460 (16.8) | 2,080 (15.8) | 2,295 (16.1) | 1,918 (13.8) | 2,300 (14.5) | 2,218 (13.9) | 2,353 (14.2) | 2,487 (15.0) | 2,331 (14.6) | 2,354 (15.8) | 2,390 (16.2) | 0.53 |
| Cardiac arrest | 770 (8.8) | 1,092 (8.3) | 1,308 (9.2) | 1,404 (10.1) | 1,626 (10.3) | 1,639 (10.3) | 1,768 (10.7) | 1,816 (10.9) | 1,852 (11.6) | 1,880 (12.6) | 1,679 (11.4) | <0.001 |
| Out-of-hospital cardiac arrest | 624 (7.2) | 869 (6.6) | 1,094 (7.7) | 1,184 (8.5) | 1,358 (8.6) | 1,365 (8.6) | 1,492 (9.0) | 1,485 (8.9) | 1,551 (9.7) | 1,555 (10.4) | 1,380 (9.4) | <0.001 |
| Treatments, *n* (%) |  |  |  |  |  |  |  |  |  |  |  |  |
| Percutaneous coronary intervention | 5,257 (60.3) | 7,915 (60.2) | 8,053 (56.7) | 7,814 (56.2) | 8,855 (55.9) | 8,934 (56.1) | 9,172 (55.3) | 8,982 (54.1) | 8,552 (53.7) | 7,928 (53.1) | 8,045 (54.6) | <0.001 |
| Coronary artery bypass grafting | 1,666 (19.1) | 2,478 (18.8) | 2,857 (20.1) | 2,774 (19.9) | 3,112 (19.6) | 3,024 (19.0) | 3,070 (18.5) | 3,038 (18.3) | 2,962 (18.6) | 2,518 (16.9) | 2,400 (16.3) | <0.001 |
| Surgical valve procedures | 528 (6.1) | 920 (7.0) | 1,148 (8.1) | 1,053 (7.6) | 1,259 (7.9) | 1,190 (7.5) | 1,260 (7.6) | 1,297 (7.8) | 1,209 (7.6) | 1,064 (7.1) | 1,100 (7.5) | 0.14 |
| Percutaneous valve procedures | 10 (0.1) | 30 (0.2) | 35 (0.2) | 67 (0.5) | 114 (0.7) | 139 (0.9) | 217 (1.3) | 297 (1.8) | 284 (1.8) | 362 (2.4) | 398 (2.7) | <0.001 |
| Outcomes |  |  |  |  |  |  |  |  |  |  |  |  |
| In-hospital mortality, *n* (%) | 2,549 (29.3) | 3,792 (28.8) | 4,180 (29.4) | 4,141 (29.8) | 4,766 (30.1) | 4,878 (30.6) | 5,085 (30.7) | 5,246 (31.6) | 5,007 (31.4) | 4,964 (33.2) | 4,799 (32.6) | <0.001 |
| Length of MCS, days, mean (SD) | 3.7 (4.4) | 3.7 (4.1) | 3.8 (4.9) | 3.8 (4.9) | 4.0 (6.5) | 4.0 (7.7) | 4.0 (5.4) | 4.2 (6.6) | 4.3 (6.4) | 4.4 (7.7) | 4.4 (6.6) | <0.001 |
| Length of hospital stay, days, mean (SD) | 29.7 (32.0) | 31.3 (37.3) | 33.0 (46.6) | 33.6 (57.2) | 31.8 (46.5) | 31.7 (43.5) | 31.6 (46.7) | 32.0 (48.1) | 31.8 (47.8) | 31.6 (44.2) | 30.9 (44.2) | 0.59 |
| Total hospitalization costs, ×10<sup>3</sup> yen, mean (SD) | 4,294 (3,040) | 4,470 (3,381) | 4,752 (4,649) | 4,892 (5,603) | 4,882 (6,076) | 4,886 (4,831) | 4,931 (5,040) | 5,043 (5,489) | 5,054 (5,188) | 5,176 (5,483) | 5,233 (5,399) | <0.001 |
| Major bleeding in a critical area or organ, *n* (%) | 32 (0.4) | 56 (0.4) | 63 (0.4) | 57 (0.4) | 109 (0.7) | 95 (0.6) | 138 (0.8) | 150 (0.9) | 126 (0.8) | 131 (0.9) | 153 (1.0) | <0.001 |
| Ischemic stroke, *n* (%) | 163 (1.9) | 269 (2.0) | 301 (2.1) | 333 (2.4) | 352 (2.2) | 382 (2.4) | 443 (2.7) | 453 (2.7) | 437 (2.7) | 372 (2.5) | 403 (2.7) | <0.001 |
| Red blood cells transfusion, ml, mean (SD) | 1,140<br>(2,309) | 1,216<br>(2,863) | 3,234<br>(55,373) | 2,596<br>(57,409) | 1,500<br>(5,350) | 1,477<br>(3,497) | 1,477<br>(2,934) | 1,573<br>(3,256) | 1,583<br>(3,130) | 1,654<br>(3,256) | 1,632<br>(3,127) | 0.14 |

Each fiscal year started on 01 April and ended on 31 March. MCS, mechanical circulatory support; SD, standard deviation

Among 41,495 patients in the ECMO group, the prevalence of ACS significantly decreased from 49.2% in 2010 to 41.5% in 2020, whereas the prevalence of valve disease significantly increased from 5.5% in 2010 to 10.5% in 2020 (**Table 3**). There was no significant change in the prevalence of heart failure and OHCA. The prevalence of patients who underwent PCI and CABG significantly decreased, whereas the prevalence of patients who underwent percutaneous valve procedures significantly increased. In-hospital mortality significantly decreased from 73.7% in 2010 to 64.1% in 2020.

**Table 3.** Patient characteristics and outcomes on an annual basis in the patients requiring ECMO.

|  | Fiscal year |  |  |  |  |  |  |  |  |  |  | *P* for trend |
| --- | --- | --- | --- | --- | --- | --- | --- | --- | --- | --- | --- | --- |
|  | 2010<br>(*n* = 1,701) | 2011<br>(*n* = 2,572) | 2012<br>(*n* = 3,148) | 2013<br>(*n* = 3,262) | 2014<br>(*n* = 3,872) | 2015<br>(*n* = 4,048) | 2016<br>(*n* = 4,406) | 2017<br>(*n* = 4,724) | 2018<br>(*n* = 4,719) | 2019<br>(*n* = 4,681) | 2020<br>(*n* = 4,362) |  |
| Age, years, mean (SD) | 64 (15) | 63 (16) | 63 (16) | 63 (16) | 63 (16) | 64 (16) | 64 (16) | 65 (16) | 65 (16) | 65 (16) | 65 (16) | 0.002 |
| Men, *n* (%) | 1,245 (73.2) | 1,880 (73.1) | 2,241 (71.2) | 2,395 (73.4) | 2,804 (72.4) | 2,897 (71.6) | 3,191 (72.4) | 3,428 (72.6) | 3,387 (71.8) | 3,332 (71.2) | 3,184 (73.0) | 0.41 |
| Body mass index on the day of admission, kg/m<sup>2</sup>, mean (SD) | 23.7 (4.0) | 23.6 (4.0) | 23.6 (4.2) | 23.7 (4.2) | 23.8 (4.4) | 23.9 (4.5) | 24.1 (9.8) | 23.9 (4.5) | 23.9 (5.0) | 24.0 (4.9) | 24.1 (4.7) | 0.001 |
| Primary diagnosis, *n* (%) |  |  |  |  |  |  |  |  |  |  |  |  |
| Acute coronary syndrome | 837 (49.2) | 1,254 (48.8) | 1,451 (46.1) | 1,519 (46.6) | 1,832 (47.3) | 1,861 (46.0) | 1,980 (44.9) | 2,021 (42.8) | 2,065 (43.8) | 1,941 (41.5) | 1,811 (41.5) | <0.001 |
| Valve disease | 93 (5.5) | 149 (5.8) | 201 (6.4) | 196 (6.0) | 294 (7.6) | 297 (7.3) | 384 (8.7) | 436 (9.2) | 419 (8.9) | 448 (9.6) | 458 (10.5) | <0.001 |
| Heart failure | 181 (10.6) | 290 (11.3) | 321 (10.2) | 330 (10.1) | 407 (10.5) | 405 (10.0) | 453 (10.3) | 507 (10.7) | 481 (10.2) | 509 (10.9) | 478 (11.0) | 0.60 |
| Cardiac arrest | 532 (31.3) | 721 (28.0) | 904 (28.7) | 948 (29.1) | 1,121 (29.0) | 1,165 (28.8) | 1,258 (28.6) | 1,315 (27.8) | 1,355 (28.7) | 1,377 (29.4) | 1,235 (28.3) | 0.38 |
| Out-of-hospital cardiac arrest | 420 (24.7) | 559 (21.7) | 744 (23.6) | 783 (24.0) | 924 (23.9) | 966 (23.9) | 1,045 (23.7) | 1,078 (22.8) | 1,134 (24.0) | 1,139 (24.3) | 1,020 (23.4) | 0.61 |
| Treatments, *n* (%) |  |  |  |  |  |  |  |  |  |  |  |  |
| Percutaneous coronary intervention | 717 (42.2) | 1,085 (42.2) | 1,226 (39.0) | 1,289 (39.5) | 1,531 (39.5) | 1,627 (40.2) | 1,735 (39.4) | 1,756 (37.2) | 1,816 (38.5) | 1,800 (38.5) | 1,659 (38.0) | <0.001 |
| Coronary artery bypass grafting | 161 (9.5) | 222 (8.6) | 306 (9.7) | 343 (10.5) | 371 (9.6) | 324 (8.0) | 367 (8.3) | 449 (9.5) | 410 (8.7) | 359 (7.7) | 323 (7.4) | <0.001 |
| Surgical valve procedures | 90 (5.3) | 141 (5.5) | 186 (5.9) | 188 (5.8) | 243 (6.3) | 222 (5.5) | 254 (5.8) | 273 (5.8) | 265 (5.6) | 234 (5.0) | 233 (5.3) | 0.21 |
| Percutaneous valve procedures | 1 (0.1) | 12 (0.5) | 12 (0.4) | 20 (0.6) | 66 (1.7) | 81 (2.0) | 133 (3.0) | 164 (3.5) | 168 (3.6) | 211 (4.5) | 208 (4.8) | <0.001 |
| Outcomes |  |  |  |  |  |  |  |  |  |  |  |  |
| In-hospital mortality, *n* (%) | 1,254 (73.7) | 1,890 (73.5) | 2,233 (70.9) | 2,315 (71.0) | 2,707 (69.9) | 2,831 (69.9) | 2,985 (67.7) | 3,156 (66.8) | 3,106 (65.8) | 3,064 (65.5) | 2,797 (64.1) | <0.001 |
| Length of MCS, days, mean (SD) | 4.6 (5.2) | 4.6 (5.8) | 4.7 (5.3) | 5.2 (6.2) | 5.4 (10.6) | 5.5 (13.8) | 5.3 (8.1) | 5.7 (10.2) | 5.7 (10.0) | 6.0 (12.4) | 5.8 (9.9) | <0.001 |
| Length of hospital stay, days, mean (SD) | 22.3 (32.1) | 24.5 (38.9) | 27.8 (50.9) | 31.5 (68.9) | 29.7 (57.5) | 30.0 (61.5) | 29.7 (58.7) | 30.7 (63.9) | 30.4 (67.5) | 30.3 (56.4) | 30.0 (56.5) | 0.04 |
| Total hospitalization costs, ×10<sup>3</sup> yen, mean (SD) | 4,700 (3,792) | 4,967 (4,294) | 5,386 (6,337) | 5,906 (7,693) | 6,029 (10,136) | 5,838 (7,005) | 5,834 (6,880) | 6,080 (7,707) | 5,943 (6,957) | 6,128 (7,753) | 6,238 (7,502) | 0.001 |
| Major bleeding in a critical area or organ, *n* (%) | 10 (0.6) | 25 (1.0) | 26 (0.8) | 30 (0.9) | 63 (1.6) | 56 (1.4) | 75 (1.7) | 93 (2.0) | 81 (1.7) | 81 (1.7) | 93 (2.1) | <0.001 |
| Ischemic stroke, *n* (%) | 30 (1.8) | 57 (2.2) | 57 (1.8) | 72 (2.2) | 102 (2.6) | 103 (2.5) | 141 (3.2) | 147 (3.1) | 147 (3.1) | 128 (2.7) | 129 (3.0) | <0.001 |
| Red blood cells transfusion, ml, mean (SD) | 2,650<br>(3,657) | 2,841<br>(3,658) | 7,443<br>(83,929) | 6,344<br>(108,475) | 3,311<br>(6,074) | 3,215<br>(4,505) | 3,112<br>(4,278) | 3,241<br>(4,798) | 3,130<br>(4,569) | 3,098<br>(4,556) | 3,077<br>(4,470) | 0.39 |

Each fiscal year started on 01 April and ended on 31 March. ECMO, extracorporeal membrane oxygenation; MCS, mechanical circulatory support; SD, standard deviation

Among 117,599 patients in the IABP alone group, the prevalence of ACS significantly decreased, whereas the prevalence of valve disease and OHCA significantly increased (**Supplementary Table S3**). There was no significant change in the prevalence of heart failure. The prevalence of patients who underwent PCI significantly decreased, whereas the prevalence of patients who underwent surgical and percutaneous valve procedures significantly increased. There was no significant change in in-hospital mortality. Among 1,465 patients in the Impella alone group, there was no significant change in the prevalence of primary diagnoses, OHCA, or treatments from 2017 to 2020 (**Supplementary Table S4**). There was also no significant change in in-hospital mortality.

## Discussion

To the best of our knowledge, this is the first nationwide study on annual trends in MCS use and outcomes in patients with CS requiring MCS in Japan. Among the overall patients with CS requiring MCS, there was a decreasing trend in IABP use, while there were increasing trends in Impella alone use and ECMO use from 2010 to 2020. There was an increasing trend in in-hospital mortality among the overall patients with CS requiring MCS, while there was a decreasing trend in in-hospital mortality among those requiring ECMO.

Consistent with previous studies from the United States and Europe,^14,22–24^ there was a decreasing trend in IABP use and increasing trends in Impella and ECMO use in the present study. However, the degree of those changes differed compared to the present study. A previous study using nationwide medical registries in Denmark that included 14,363 patients with CS regardless of MCS use showed that among the patients requiring MCS, there was a drastic decrease in IABP use from 90% in 2010 to 5.3% in 2017 and an increase in Impella and ECMO use from 4.5% and 5.5% in 2010 to 36% and 59% in 2017, respectively.^14^ Even when there were strong guideline recommendations prior to the downgrade, the prevalence of IABP use in patients with CS due to STEMI remained low at 20–39%.^25^ This may be partly because interventionists were not fully convinced about these recommendations based on non-randomized trials and registries, and were concerned about potential complications.^26^ The negative results from the landmark randomized controlled trial, Intra-aortic Balloon Pump in Cardiogenic Shock (IABP-SHOCK) II trial^8^, in 2012 and subsequent revisions of the guidelines may have accelerated the shift away from IABP.^27^ In contrast, the present study revealed that the IABP use remained high in 2020 in Japan. This result may be partly due to the quite high penetration of IABP in acute care hospitals in Japan, the lower profile of IABP catheters as compared to Impella and ECMO catheters, and its frequent use in elective high-risk PCI cases.^28^ In addition, prompt regulatory approval, physician training programs, generous reimbursement for Impella use, and ongoing clinical trials have led to a substantial increase in Impella use in the United States and Europe.^4,29^ However, in Japan, the Impella remains available at only a few acute care hospitals and is not indicated for high-risk PCI cases, and is limited to use in patients with CS.

In the present study, in-hospital mortality was approximately 30%, which was almost consistent with that in the previous studies focusing on the most recent decade.^22,30^ However, unlike the previous studies that showed a decreasing trend in in-hospital mortality,^22,30^ the present study showed an increasing trend in in-hospital mortality. This may be attributed to the increasing prevalence of patients requiring ECMO in the present study, who had the highest in-hospital mortality as compared to those requiring IABP alone or Impella alone. In addition, the present study included only patients with CS requiring MCS and had a high prevalence of ECMO use, while the previous studies included all patients with CS regardless of requiring MCS and who had a low prevalence of ECMO use among the patients with CS requiring MCS (26% vs. 3.2–3.9%).^22,30^ A potential reason for the increasing prevalence of patients requiring ECMO in the present study may be the increasing prevalence of OHCA, in contrast to the decreasing prevalence of OHCA in the previous study.^30^ Although in-hospital mortality in patients requiring ECMO decreased over time in the present study, further mortality reductions would be desirable to decrease in-hospital mortality in the overall patients with CS requiring MCS. One possible solution might be the concomitant use of an IABP or Impella with ECMO to achieve left ventricular unloading.^31^

Our results included some important clinical implications. First, the downgrade of the guideline recommendations for IABP use and approval of the Impella changed the pattern of MCS use to some degree, but not drastically in Japan, where the Impella is still in the process of widespread use. The use of the Impella is likely to become more widespread with a decrease in IABP use, and studies are warranted to capture the trends in the pattern of MCS use and outcomes. Second, when limited to patients requiring ECMO, in-hospital mortality decreased over time, but was still approximately twice as high as that in the overall patients with CS requiring MCS. In order to decrease in-hospital mortality in the overall patients with CS requiring MCS, further mortality reductions would be warranted in patients requiring ECMO. The concomitant use of an IABP or Impella with ECMO might be a key to achieve this, and further investigation is required.

The present study had several strengths. First, the present study was based on one of the largest databases, which covered approximately 90% of all tertiary emergency hospitals in Japan. Second, our database included data before and after the change in the indication for IABP and the approval of the Impella.

The present study had several limitations. First, our definition of CS depended on the administrative data on MCS use during the hospitalization in patients with the primary diagnosis of cardiovascular disease because the database does not contain detailed clinical information such as the vital signs and laboratory tests. Therefore, misclassifications may have led to a bias in our study. Second, we could not identify the patients who received ECMO with concomitant IABP or Impella use because the simultaneous cost calculation of MCS was not allowed in Japan. Finally, the incidence of major bleeding events in the present study was considerably lower than in the previous studies.^4,22,24^ Given that the sensitivity of the diagnosis might have been low in our database, there was a possibility of underreporting major bleeding events.

## Conclusions

The present study using a nationwide inpatient administrative database showed significant annual changes in the patterns of MCS use and clinical outcomes in patients with CS requiring MCS. In the overall patients with CS requiring MCS, the annual prevalence of IABP alone use decreased, albeit the most frequent, while those of Impella alone and ECMO use increased. In-hospital mortality increased in the overall patients with CS requiring MCS, but decreased in those requiring ECMO.

## Data Availability

The datasets analyzed during the current study are not publicly available owing to contracts with the hospitals providing the data to the database.

## Declarations

### Ethics approval and consent to participate

This study was performed in accordance with the amended Declaration of Helsinki, and the Institutional Review Board of The University of Tokyo approved this study (approval number: 3501-(3); 25 December 2017).

### Consent for publication

The review board waived the requirement for informed consent because of the anonymous nature of the data. No information describing the individual patients, hospitals, or treating physicians was obtained.

### Authors’ contributions

**Yuji Nishimoto**: Conceptualization, Software, Formal analysis, Investigation, Writing - Original Draft; **Hiroyuki Ohbe**: Conceptualization, Methodology, Software, Formal analysis, Investigation, Writing - Review and Editing; **Hiroki Matsui**: Software, Formal analysis, Investigation, Data Curation; **Jun Nakata**: Conceptualization, Methodology, Investigation, Writing - Review and Editing, Supervision; **Toru Takiguchi**: Conceptualization, Methodology, Investigation, Writing - Review and Editing, Supervision; **Mikio Nakajima**: Conceptualization, Investigation, Writing - Review and Editing, Supervision; **Yusuke Sasabuchi**: Conceptualization, Investigation, Writing - Review and Editing, Supervision; **Yukihito Sato:** Software, Investigation, Resources, Supervision; **Tetsuya Watanabe**: Software, Investigation, Resources, Supervision; **Takahisa Yamada**: Software, Investigation, Resources, Supervision; **Masatake Fukunami**: Software, Investigation, Resources, Supervision; **Hideo Yasunaga**: Writing - Review and Editing, Supervision. All authors read and approved the final manuscript.

## Acknowledgments

We would like to express our gratitude to Mr. John Martin for his grammatical assistance.

## References

1. van Diepen S, Katz JN, Albert NM, Henry TD, Jacobs AK, Kapur NK, Kilic A, Menon V, Ohman EM, Sweitzer NK, Thiele H, Washam JB, Cohen MG. Contemporary Management of Cardiogenic Shock: A Scientific Statement From the American Heart Association. Circulation. 2017;136:e232–268.

2. Baran DA, Grines CL, Bailey S, Burkhoff D, Hall SA, Henry TD, Hollenberg SM, Kapur NK, O’Neill W, Ornato JP, Stelling K, Thiele H, Diepen S, Naidu SS. SCAI clinical expert consensus statement on the classification of cardiogenic shock: This document was endorsed by the American College of Cardiology (ACC), the American Heart Association (AHA), the Society of Critical Care Medicine (SCCM), and the Society of Thoracic Surgeons (STS) in April 2019. Catheter Cardiovasc Interv. 2019;94:29–37.

3. Kolte D, Khera S, Aronow WS, Mujib M, Palaniswamy C, Sule S, Jain D, Gotsis W, Ahmed A, Frishman WH, Fonarow GC. Trends in Incidence, Management, and Outcomes of Cardiogenic Shock Complicating ST-Elevation Myocardial Infarction in the United States. JAHA. 2014;3:e000590.

4. Helgestad OKL, Josiassen J, Hassager C, Jensen LO, Holmvang L, Sørensen A, Frydland M, Lassen AT, Udesen NLJ, Schmidt H, Ravn HB, Møller JE. Temporal trends in incidence and patient characteristics in cardiogenic shock following acute myocardial infarction from 2010 to 2017: a Danish cohort study. Eur J Heart Fail. 2019;21:1370–1378.

5. Berg DD, Bohula EA, van Diepen S, Katz JN, Alviar CL, Baird-Zars VM, Barnett CF, Barsness GW, Burke JA, Cremer PC, Cruz J, Daniels LB, DeFilippis AP, Haleem A, Hollenberg SM, Horowitz JM, Keller N, Kontos MC, Lawler PR, Menon V, Metkus TS, Ng J, Orgel R, Overgaard CB, Park J-G, Phreaner N, Roswell RO, Schulman SP, Jeffrey Snell R, Solomon MA, Ternus B, Tymchak W, Vikram F, Morrow DA. Epidemiology of Shock in Contemporary Cardiac Intensive Care Units: Data From the Critical Care Cardiology Trials Network Registry. Circ: Cardiovascular Quality and Outcomes. 2019;12:e005618.

6. Osman M, Syed M, Patibandla S, Sulaiman S, Kheiri B, Shah MK, Bianco C, Balla S, Patel B. Fifteen-Year Trends in Incidence of Cardiogenic Shock Hospitalization and In-Hospital Mortality in the United States. JAHA. 2021;10:e021061.

7. Patel MR, Smalling RW, Thiele H, Barnhart HX, Zhou Y, Chandra P, Chew D, Cohen M, French J, Perera D, Ohman EM. Intra-aortic balloon counterpulsation and infarct size in patients with acute anterior myocardial infarction without shock: the CRISP AMI randomized trial. JAMA. 2011;306:1329–1337.

8. Thiele H, Zeymer U, Neumann F-J, Ferenc M, Olbrich H-G, Hausleiter J, Richardt G, Hennersdorf M, Empen K, Fuernau G, Desch S, Eitel I, Hambrecht R, Fuhrmann J, Böhm M, Ebelt H, Schneider S, Schuler G, Werdan K, IABP-SHOCK II Trial Investigators. Intraaortic balloon support for myocardial infarction with cardiogenic shock. N Engl J Med. 2012;367:1287–1296.

9. Ahmad Y, Sen S, Shun-Shin MJ, Ouyang J, Finegold JA, Al-Lamee RK, Davies JER, Cole GD, Francis DP. Intra-aortic Balloon Pump Therapy for Acute Myocardial Infarction: A Meta-analysis. JAMA Intern Med. 2015;175:931–939.

10. O’Gara PT, Kushner FG, Ascheim DD, Casey DE, Chung MK, de Lemos JA, Ettinger SM, Fang JC, Fesmire FM, Franklin BA, Granger CB, Krumholz HM, Linderbaum JA, Morrow DA, Newby LK, Ornato JP, Ou N, Radford MJ, Tamis-Holland JE, Tommaso CL, Tracy CM, Woo YJ, Zhao DX, Anderson JL, Jacobs AK, Halperin JL, Albert NM, Brindis RG, Creager MA, DeMets D, Guyton RA, Hochman JS, Kovacs RJ, Kushner FG, Ohman EM, Stevenson WG, Yancy CW, American College of Cardiology Foundation/American Heart Association Task Force on Practice Guidelines. 2013 ACCF/AHA guideline for the management of ST-elevation myocardial infarction: a report of the American College of Cardiology Foundation/American Heart Association Task Force on Practice Guidelines. Circulation. 2013;127:e362–425.

11. Authors/Task Force members, Windecker S, Kolh P, Alfonso F, Collet J-P, Cremer J, Falk V, Filippatos G, Hamm C, Head SJ, Jüni P, Kappetein AP, Kastrati A, Knuuti J, Landmesser U, Laufer G, Neumann F-J, Richter DJ, Schauerte P, Sousa Uva M, Stefanini GG, Taggart DP, Torracca L, Valgimigli M, Wijns W, Witkowski A. 2014 ESC/EACTS Guidelines on myocardial revascularization: The Task Force on Myocardial Revascularization of the European Society of Cardiology (ESC) and the European Association for Cardio-Thoracic Surgery (EACTS)Developed with the special contribution of the European Association of Percutaneous Cardiovascular Interventions (EAPCI). Eur Heart J. 2014;35:2541–2619.

12. Kimura K, Kimura T, Ishihara M, Nakagawa Y, Nakao K, Miyauchi K, Sakamoto T, Tsujita K, Hagiwara N, Miyazaki S, Ako J, Arai H, Ishii H, Origuchi H, Shimizu W, Takemura H, Tahara Y, Morino Y, Iino K, Itoh T, Iwanaga Y, Uchida K, Endo H, Kongoji K, Sakamoto K, Shiomi H, Shimohama T, Suzuki A, Takahashi J, Takeuchi I, Tanaka A, Tamura T, Nakashima T, Noguchi T, Fukamachi D, Mizuno T, Yamaguchi J, Yodogawa K, Kosuge M, Kohsaka S, Yoshino H, Yasuda S, Shimokawa H, Hirayama A, Akasaka T, Haze K, Ogawa H, Tsutsui H, Yamazaki T, on behalf of the Japanese Circulation Society Joint Working Group. JCS 2018 Guideline on Diagnosis and Treatment of Acute Coronary Syndrome. Circ J. 2019;83:1085–1196.

13. Lauridsen MD, Rørth R, Lindholm MG, Kjaergaard J, Schmidt M, Møller JE, Hassager C, Torp-Pedersen C, Gislason G, Køber L, Fosbøl EL. Trends in first-time hospitalization, management, and short-term mortality in acute myocardial infarction–related cardiogenic shock from 2005 to 2017: A nationwide cohort study. American Heart Journal. 2020;229:127–137.

14. Petersen LT, Riddersholm S, Andersen DC, Polcwiartek C, Lee CJ-Y, Lauridsen MD, Fosbøl E, Christiansen CF, Pareek M, Søgaard P, Torp-Pedersen C, Rasmussen BS, Kragholm KH. Temporal trends in patient characteristics, presumed causes, and outcomes following cardiogenic shock between 2005 and 2017: a Danish registry-based cohort study. European Heart Journal Acute Cardiovascular Care. 2021;10:1074–1083.

15. Inohara T, Miyata H, Ueda I, Maekawa Y, Fukuda K, Kohsaka S. Use of Intra-aortic Balloon Pump in a Japanese Multicenter Percutaneous Coronary Intervention Registry. JAMA Intern Med. 2015;175:1980–1982.

16. Deedwania P, Acharya T. Is Increased Use of Mechanical Circulatory Support Devices Justified? A Cause for Concern. JAMA Intern Med. 2015;175:1982–1983.

17. Benchimol EI, Smeeth L, Guttmann A, Harron K, Moher D, Petersen I, Sørensen HT, von Elm E, Langan SM, RECORD Working Committee. The REporting of studies Conducted using Observational Routinely-collected health Data (RECORD) statement. PLoS Med. 2015;12:e1001885.

18. Yasunaga H. Real World Data in Japan: Chapter II The Diagnosis Procedure Combination Database. ACE. 2019;1:76–79.

19. Yamana H, Moriwaki M, Horiguchi H, Kodan M, Fushimi K, Yasunaga H. Validity of diagnoses, procedures, and laboratory data in Japanese administrative data. J Epidemiol. 2017;27:476–482.

20. Shigematsu K, Nakano H, Watanabe Y. The eye response test alone is sufficient to predict stroke outcome--reintroduction of Japan Coma Scale: a cohort study. BMJ Open. 2013;3:e002736.

21. Schulman S, Angerås U, Bergqvist D, Eriksson B, Lassen MR, Fisher W, Subcommittee on Control of Anticoagulation of the Scientific and Standardization Committee of the International Society on Thrombosis and Haemostasis. Definition of major bleeding in clinical investigations of antihemostatic medicinal products in surgical patients. J Thromb Haemost. 2010;8:202–204.

22. Strom JB, Zhao Y, Shen C, Chung M, Pinto DS, Popma JJ, Yeh RW. National trends, predictors of use, and in-hospital outcomes in mechanical circulatory support for cardiogenic shock. EuroIntervention. 2018;13:2152–2159.

23. Amin AP, Spertus JA, Curtis JP, Desai N, Masoudi FA, Bach RG, McNeely C, Al-Badarin F, House JA, Kulkarni H, Rao SV. The Evolving Landscape of Impella Use in the United States Among Patients Undergoing Percutaneous Coronary Intervention With Mechanical Circulatory Support. Circulation. 2020;141:273–284.

24. Helgestad OKL, Josiassen J, Hassager C, Jensen LO, Holmvang L, Udesen NLJ, Schmidt H, Berg Ravn H, Moller JE. Contemporary trends in use of mechanical circulatory support in patients with acute MI and cardiogenic shock. Open Heart. 2020;7:e001214.

25. Unverzagt S, Buerke M, de Waha A, Haerting J, Pietzner D, Seyfarth M, Thiele H, Werdan K, Zeymer U, Prondzinsky R. Intra-aortic balloon pump counterpulsation (IABP) for myocardial infarction complicated by cardiogenic shock. Cochrane Database of Systematic Reviews. 2015;2015:CD007398.

26. Rathod KS, Koganti S, Iqbal MB, Jain AK, Kalra SS, Astroulakis Z, Lim P, Rakhit R, Dalby MC, Lockie T, Malik IS, Knight CJ, Whitbread M, Mathur A, Redwood S, MacCarthy PA, Sirker A, O’Mahony C, Wragg A, Jones DA. Contemporary trends in cardiogenic shock: Incidence, intra-aortic balloon pump utilisation and outcomes from the London Heart Attack Group. European Heart Journal: Acute Cardiovascular Care. 2018;7:16–27.

27. Nevzorov R, Daum A, Jafari J, Yosefy C, Gallego-Colon E. Impact of the Change in ESC Guidelines on Clinical Characteristics and Outcomes of Cardiogenic Shock Patients Receiving IABP Therapy. Cardiovasc Revasc Med. 2020;21:46–51.

28. Muramatsu T, Inohara T, Kohsaka S, Yamaji K, Ishii H, Shinke T, Toriya T, Yoshiki Y, Ozaki Y, Ando H, Amano T, Nakamura M, Ikari Y. Mechanical circulatory support devices for elective percutaneous coronary interventions: novel insights from the Japanese nationwide J-PCI registry. European Heart Journal Open. 2022;2:oeac041.

29. Khera R, Cram P, Lu X, Vyas A, Gerke A, Rosenthal GE, Horwitz PA, Girotra S. Trends in the Use of Percutaneous Ventricular Assist Devices: Analysis of National Inpatient Sample Data, 2007 Through 2012. JAMA Intern Med. 2015;175:941.

30. Jentzer JC, Ahmed AM, Vallabhajosyula S, Burstein B, Tabi M, Barsness GW, Murphy JG, Best PJ, Bell MR. Shock in the cardiac intensive care unit: Changes in epidemiology and prognosis over time. American Heart Journal. 2021;232:94–104.

31. Russo JJ, Aleksova N, Pitcher I, Couture E, Parlow S, Faraz M, Visintini S, Simard T, Di Santo P, Mathew R, So DY, Takeda K, Garan AR, Karmpaliotis D, Takayama H, Kirtane AJ, Hibbert B. Left Ventricular Unloading During Extracorporeal Membrane Oxygenation in Patients With Cardiogenic Shock. Journal of the American College of Cardiology. 2019;73:654–662.

